# Rapid tests as a practical alternative to slide agglutination for the confirmation of *V. cholerae* O1

**DOI:** 10.1101/2025.03.19.25324236

**Authors:** Piyash Bhattacharjee, Sonia T Hegde, Ashraful Islam Khan, Imrul Kayes Nabil, Md. Naiem Hossain, Tahira Ahmed Rashmi, Mokibul Hassan Afrad, Md. Taufiqul Islam, Mohammad Ashraful Amin, Zahid Hasan Khan, Taufiqur Rahman Bhuiyan, Andrew S Azman, Firdausi Qadri

## Abstract

**Objectives:** Slide agglutination is a crucial step for confirming cholera culture by determining serogroup/serotype. Rapid diagnostic tests (RDTs), typically used on stool, may provide a practical alternative, as they are easy to use, store and require minimal training. This study evaluates the concordance of *V. cholerae* O1/O139 detection from presumptive colonies by slide agglutination with RDTs.

**Methods:** Patients (≥1 year) with acute watery diarrhea at the icddr,b Dhaka hospital were enrolled. Stool samples cultured on Thiosulfate-Citrate-Bile Salts-Sucrose media, and presumptive colonies were sub-cultured on Gelatin Agar. Isolates were then tested by slide agglutination and four commercial cholera RDTs.

**Results:** From 4-February-2024 through 31-January-2025, 1,331 patients with acute watery diarrhea were enrolled, 952 had presumptive colonies and 408 (31%) were culture-confirmed as *V. cholerae* O1. We tested 952, 505, 505, and 467 presumptive colonies with Cholkit, SD-Bioline, Crystal VC O1/O139, and Crystal VC O1 RDT kits, respectively. RDTs showed near-perfect concordance with slide agglutination. Using slide agglutination as the reference, the sensitivity of the RDT kits ranged from 99.5-100% and the specificity from 99-99.8%. No *V. cholerae* O139 was detected by slide agglutination or PCR.

**Conclusions:** RDTs offer a practical, and potentially easier alternative to slide agglutination of presumptive *cholerae* colonies within typical cholera culture protocols. This may help to provide a pathway to quick confirmation of outbreaks in settings where lab facilities and reagents may be limited.

## Introduction

Cholera continues as a global public health concern, causing significant mortality and morbidity in addition to economic strains in communities lacking safe water and sanitation. While not considered useful for individual-level clinical decision-making, laboratory confirmation of suspected cholera cases is critical for declaring outbreaks, tracking trends in incidence, and monitoring antimicrobial resistance patterns of the bacteria [1]. Both Polymerase Chain Reaction (PCR) and culture are considered gold-standard methods to confirm *Vibrio cholerae* O1/O139, though PCR is not widely used in settings where cholera transmission occurs.

Stool culture-based diagnosis of *Vibrio cholerae* involves first growing the bacteria in a selective media and then confirming the serogroup and/or serotype of suspected bacterial colonies by conducting slide agglutination with antisera [2,3]. The antisera needed to confirm cholera is expensive, has a short shelf life – especially compared to the recurrent frequency of cholera in some settings – and requires skilled personnel [4]. Lack of antisera has been reported as a reason for delayed confirmation of outbreaks in several settings over the past decade. Due to the often-weak visual signal, interpretation of agglutination tests can vary from person-to-person and can be influenced by uncorrected near-vision impairments due to a lack of ocular healthcare, which is common in low-and-middle-income settings where cholera usually occurs [5].

Commercially available lateral flow immunoassay rapid diagnostic tests (RDTs) for cholera, which are based on the detection of the O-antigen of the lipopolysaccharide antigen, may offer an alternative, efficient, and perhaps more robust method to confirm *V. cholerae* O1 from suspected bacterial colonies during culture [6,7]. Here we compare the use of RDTs with slide agglutination to confirm suspected colonies as *V cholerae* O1/O139 as part of culture confirmation.

## Methods

As part of a larger evaluation of RDT performance, we recruited suspected cholera cases (patients experiencing acute watery diarrhea with three or more non-bloody, loose stools and symptoms in the preceding 24 hours prior to the hospital visit) from the icddr,b diarrhea hospital in Dhaka, Bangladesh, from 4-February-2024 through 31-January-2025. After providing informed written consent/assent, study staff administrated a short questionnaire and collected a stool sample from each case.

Within one hour of collection, stool samples from each case were cultured overnight on Thiosulfate-Citrate-Bile Salts-Sucrose culture media with suspected colonies then subcultured on gelatin agar culture media. Suspected colonies from the gelatin agar culture plate were then tested by slide agglutination with both *V. cholerae* O1 and O139 anti-sera. We put 2-3 presumptive colonies from the same gelatin agar culture plate in the diluents provided with the testing kits and then conducted four different RDTs: Cholkit (Incepta, Bangladesh), SD-Bioline (Abbott, Republic of Korea), Crystal VC O1/O139 (Arkay, India), and Crystal VC O1 (Arkay, India). Cholkit and Crystal VC O1 kits detect *V. cholerae* O1, and SD-Bioline and Crystal VC O1/O139 detect both *V. cholerae* O1 and O139. Additionally, PCR tests for *V. cholerae* O1 and O139 were performed for further molecular-level confirmation on stored stool specimens, one week after sample collection [8]. This study was started only using CholKit RDTs to compare to agglutination, but after promising preliminary results, we expanded this to all kits; thus, the numbers of tests performed for each test differed.

## Results

We recruited 1,331 suspected cholera cases from icddr,b hospital in Dhaka from 4 February 2024 through 31 January 2025, including 36% severely dehydrated [9], 40% under 5 years old, and 47% female. Among the samples, 952 had suspected *V. cholerae* colonies, and 408 (31%) were culture-positive for *V. cholerae* O1 by culture, including slide agglutination. No samples were positive for *V. cholerae O139*.

We tested 952, 505, 505, and 467 suspected *V. cholerae* colonies using Cholkit, SD-Bioline, Crystal VC O1/O139, and Crystal VC O1 RDT kits, respectively. We found nearly perfect concordance of the slide agglutination results with all four RDTs, with only seven isolates having discordant results (Table 1). Considering slide agglutination as the reference standard for confirmation of *V. cholerae* O1 among suspected colonies, the sensitivity of tests was 99.5–100%, and the specificity ranged from 99–99.8% across tests (Table 2).

**Table 1.**
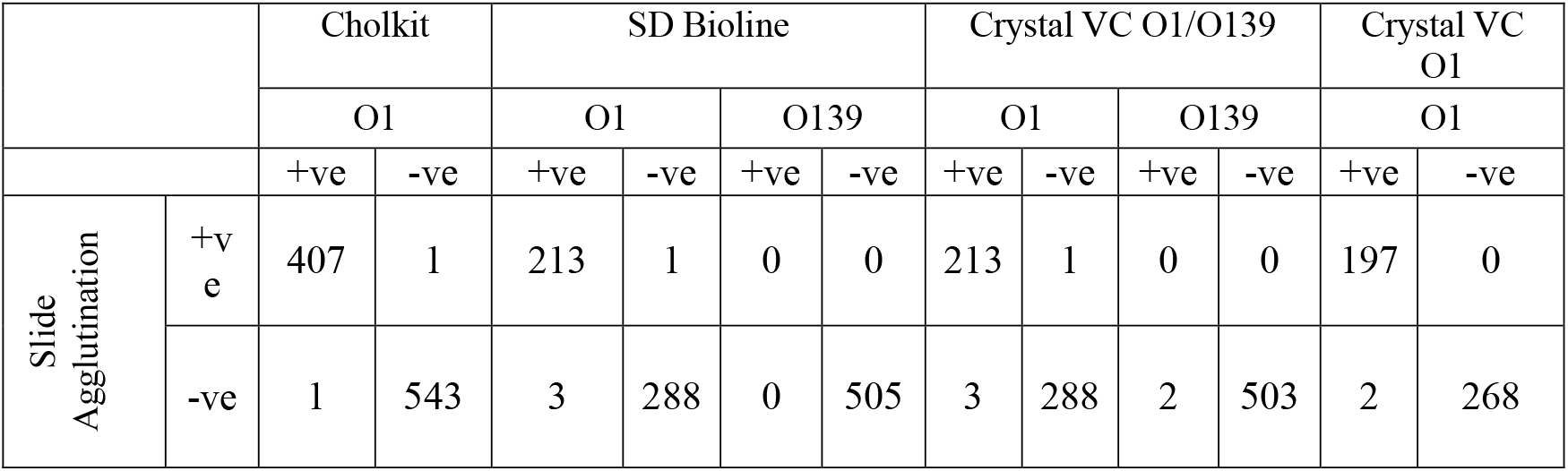
Diagnostics based on slide agglutination and RDT test results in terms of detection of *Vibrio cholerae* O1. +ve: positive; -ve: negative.

|  |  | Cholkit |  | SD Bioline |  |  |  | Crystal VC O1/O139 |  |  |  | Crystal VC O1 |  |
| --- | --- | --- | --- | --- | --- | --- | --- | --- | --- | --- | --- | --- | --- |
|  |  | O1 |  | O1 |  | O139 |  | O1 |  | O139 |  | O1 |  |
|  |  | +ve | -ve | +ve | -ve | +ve | -ve | +ve | -ve | +ve | -ve | +ve | -ve |
| Slide Agglutination | +ve | 407 | 1 | 213 | 1 | 0 | 0 | 213 | 1 | 0 | 0 | 197 | 0 |
|  | -ve | 1 | 543 | 3 | 288 | 0 | 505 | 3 | 288 | 2 | 503 | 2 | 268 |

**Table 2.** Sensitivity and specificity of RDTs for detection of *Vibrio cholerae* O1 within suspected cultured colonies as compared to slide agglutination.

|  | Compared to slide agglutination |  |
| --- | --- | --- |
|  | Sensitivity | Specificity |
| *Cholkit* | 99.8% | 99.8% |
| *SD Bioline* | 99.5% | 99.0% |
| *Crystal VC O1/O139* | 99.5% | 99.0% |
| *Crystal VC O1* | 100% | 99.3% |

Two isolates were positive for *V. cholerae* O139 by Crystal VC O1/O139 with both testing negative by culture/slide agglutination and PCR. We had only one instance of a slide agglutination positive sample being negative on RDT (CholKit) and this sample was *V. cholerae* O1 positive by PCR. Five samples were slide-agglutination negative and RDT positive (SD-Bioline, Crystal VC O1/O139, Crystal VC O1), with two samples testing negative and the other three testing positive by PCR.

## Discussion

Our study found nearly perfect concordance between slide agglutination for *V. cholerae* O1 and four commercially available cholera rapid diagnostic tests. These results suggest that RDTs could be used as an alternative to slide agglutination for confirming cholera when antisera and/or well-trained laboratory staff are unavailable.

While not often reported in the formal literature, lack of culture facilities and maintenance of usable stocks of antiserum in low-resource laboratories is challenging and has led to delayed outbreak confirmations in several countries. The cost of antisera, if purchased commercially, is roughly 4 USD per sample with a shelf-life under cold-chain of two years. In comparison, RDTs typically cost around 2 USD per test with a similar shelf life at ambient temperatures, making them easier to store and transport and not requiring laboratory infrastructure [10,11].

While we aimed to show the concordance of slide agglutination and RDT confirmation using highly trained laboratory technicians, these findings additionally suggest that RDTs may have superior practical performance, especially in laboratories where agglutination tests are not routinely performed. Immunochromatographic tests, such as the RDTs used in this study, provide clear color bands to indicate a positive test and a control band to indicate when the test is not working as expected (Figure S1). In contrast, slide agglutination relies on detecting visible clumping or lower intensities of agglutination, depending on the number of bacteria present, something which can be subjective and maybe quite hard to interpret, particularly when technicians have minor vision impairments (Figure S1).

Our results show that RDTs can be used as an alternative to slide agglutination for the confirmation of *V. cholerae* O1 and may even lead to more consistent and reliable results. As RDTs become more widely available across cholera affected countries [12, 13], stockouts of antisera should no longer inhibit confirmation of cholera outbreaks.

## Transparency declaration

### Potential conflict of interest

All authors declare that they have no conflicts of interest.

### Financial report

This work was supported by a grant from the Bill and Melinda Gates Foundation (INV-044856).

## Data Availability

Non-identifiable data produced in the study are available upon reasonable request to the authors.

## Author contributions

FQ and ASA conceptualized the study. PB, IKN, MNH, TA, MHA participated in development and methodology. PB, IKN, MNH, TA, MHA, AIK, TI, ZHK, MAA participated in data collection. ASA, TRB, PB, STH participated in data analysis and interpretation. PB, TRB, STH, ASA participated in writing the first draft of the manuscript. All authors participated in revision of the manuscript.

## Acknowledgment

icddr,b is also grateful to the Government of Bangladesh and Canada for providing core/unrestricted support.

---

**Figure S1.**
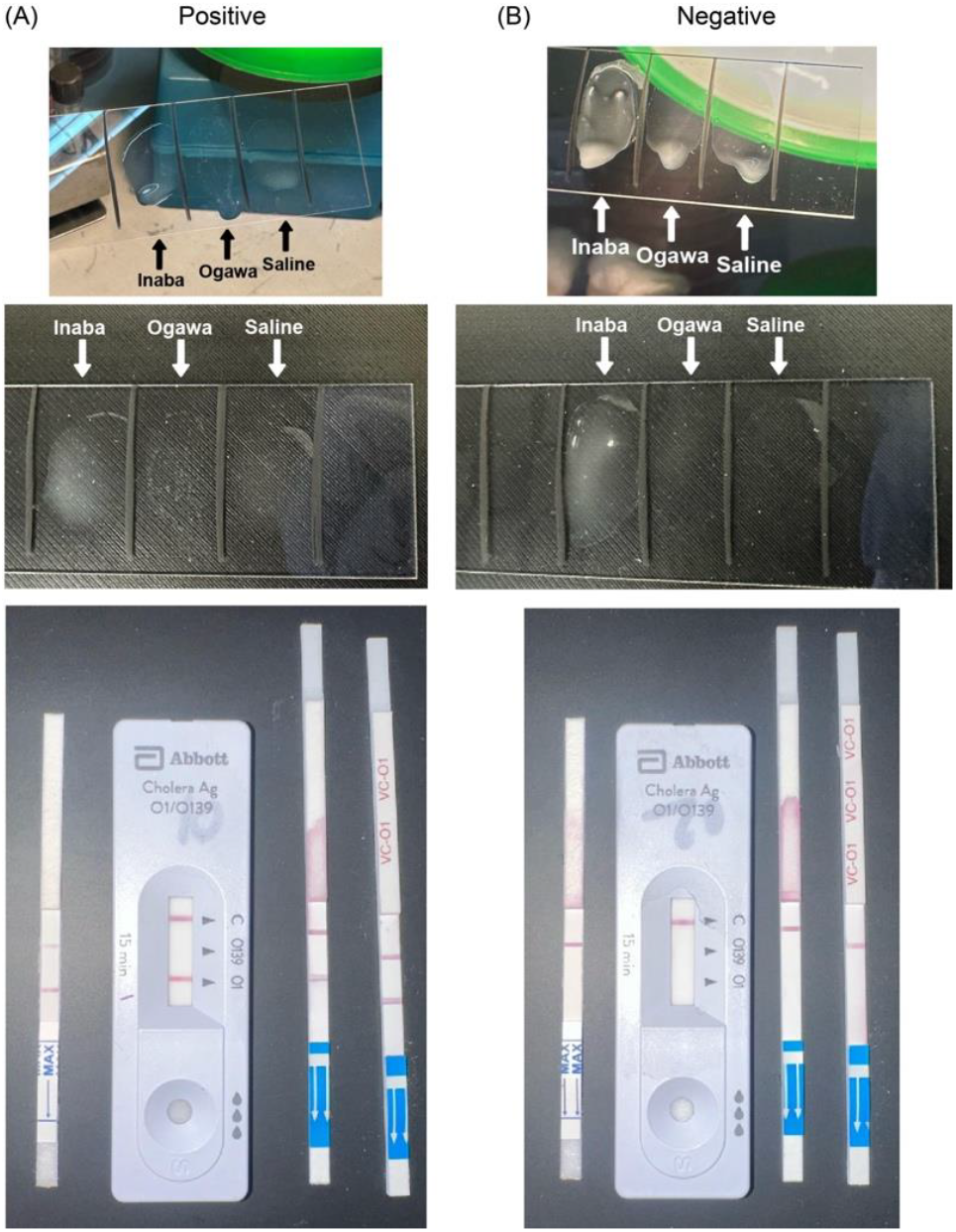
Illustration of a positive (left) and negative (right) sample tested by slide agglutination (top two photos) and RDT (bottom photos).

## Notes

### Competing Interest Statement

The authors have declared no competing interest.

### Author Declarations

This study was reviewed and approved by the icddr,b Ethical Review Committee (PR-23069) and the Johns Hopkins Bloomberg School of Public Health Institutional Review Board (IRB No. 25419)

## References

1. https://www.gtfcc.org/resources/public-health-surveillance-for-cholera/

2. Azman, Andrew S., Iza Ciglenecki, and Francisco J. Luquero. “Putting rapid tests to work in surveillance and control of cholera.” Clinical Microbiology and Infection 28, no. 2 (2022): 152–154.

3. National Center for Infectious Diseases (U.S) “Laboratory methods for the diagnosis of Vibrio cholerae” (1994)

4. Matias, Wilfredo R., Fabrice E. Julceus, Cademil Abelard, Leslie M. Mayo-Smith, Molly F. Franke, Jason B. Harris, and Louise C. Ivers. “Laboratory evaluation of immunochromatographic rapid diagnostic tests for cholera in Haiti.” PLoS One 12, no. 11 (2017): e0186710.

5. Burton, Matthew J., Jacqueline Ramke, Ana Patricia Marques, Rupert RA Bourne, Nathan Congdon, Iain Jones, Brandon AM Ah Tong et al. “The lancet global health commission on global eye health: vision beyond 2020.” The Lancet Global Health 9, no. 4 (2021): e489–e551.

6. Sayeed, Md Abu, Kamrul Islam, Motaher Hossain, Noor Jahan Akter, Md Nur Alam, Nishat Sultana, Farhana Khanam et al. “Development of a new dipstick (Cholkit) for rapid detection of Vibrio cholerae O1 in acute watery diarrheal stools.” PLoS neglected tropical diseases 12, no. 3 (2018): e0006286.

7. Muzembo, Basilua Andre, Kei Kitahara, Anusuya Debnath, Keinosuke Okamoto, and Shin-Ichi Miyoshi. “Accuracy of cholera rapid diagnostic tests: a systematic review and meta-analysis.” Clinical Microbiology and Infection 28, no. 2 (2022): 155–162.

8. Hoshino, Katsuaki, Shinji Yamasaki, Asish K. Mukhopadhyay, Soumen Chakraborty, Arnab Basu, Sujit K. Bhattacharya, G. Balakrish Nair, Toshio Shimada, and Yoshifumi Takeda. “Development and evaluation of a multiplex PCR assay for rapid detection of toxigenic Vibrio cholerae O1 and O139.” FEMS Immunology & Medical Microbiology 20, no. 3 (1998): 201–207.

9. Levine AC, Glavis-Bloom J, Modi P, et al. External validation of the DHAKA score and comparison with the current IMCI algorithm for the assessment of dehydration in children with diarrhoea: a prospective cohort study. Lancet Glob Health. 2016;4(10):e744–e751. doi:10.1016/S2214-109X(16)30150-4.

10. Chibwe, Innocent, Watipaso Kasambara, Mathews Kagoli, Harry Milala, Charity Gondwe, and Andrew S. Azman. “Field evaluation of Cholkit rapid diagnostic test for Vibrio cholerae O1 during a cholera outbreak in Malawi, 2018.” In Open Forum Infectious Diseases, vol. 7, no. 11, p. ofaa493. US: Oxford University Press, 2020.

11. Ramamurthy, Thandavarayan, Bhabatosh Das, Subhra Chakraborty, Asish K. Mukhopadhyay, and David A. Sack. “Diagnostic techniques for rapid detection of Vibrio cholerae O1/O139.” Vaccine 38 (2020): A73–A82.

12. Islam, Md Taufiqul, Ashraful Islam Khan, Md Abu Sayeed, Jakia Amin, Kamrul Islam, Nur Alam, Nishat Sultana et al. “Field evaluation of a locally produced rapid diagnostic test for early detection of cholera in Bangladesh.” PLoS neglected tropical diseases 13, no. 1 (2019): e0007124.

13. Global Task Force on Cholera Control (GTFCC), WHO. Public health surveillance for cholera (Interim guidance). 2023.

